# SARS-CoV-2 within-host and *in-vitro* genomic variability and sub-genomic RNA levels indicate differences in viral expression between clinical and *in-vitro* cohorts

**DOI:** 10.1101/2021.11.23.21266789

**Authors:** Jessica E. Agius, Jessica C. Johnson-Mackinnon, Winkie Fong, Mailie Gall, Connie Lam, Kerri M. Basile, Jen Kok, Alicia Arnott, Vitali Sintchenko, Rebecca J. Rockett

**Author notes:** **Corresponding author:** Dr Jessica Johnson-Mackinnon, Sydney Medical School, Faculty of Medicine and Health, University of Sydney, Westmead Hospital Westmead, NSW, 2145, Australia. These authors have contributed equally to this work and share first authorship (Jessica E. Agius, Jessica C. Johnson-Mackinnon). These authors have contributed equally to this work and share last authorship (Vitali Sintchenko, Rebecca J. Rockett).

## Abstract

**Background:** Low frequency intrahost single nucleotide variants (iSNVs) of Severe Acute Respiratory Syndrome Coronavirus 2 (SARS-CoV-2) have been increasingly recognised as predictive indicators of positive selection. Particularly as growing numbers of SARS-CoV-2 variants of interest (VOI) and concern (VOC) emerge. However, the dynamics of subgenomic RNA (sgRNA) expression and its impact on genomic diversity and infection outcome remain poorly understood. This study aims to investigate and quantify iSNVs and sgRNA expression in single and longitudinally sampled cohorts over the course of mild and severe SARS-CoV-2 infection benchmarked against an *in-vitro* infection model.

**Methods:** Two clinical cohorts of SARS-CoV-2 positive cases in New South Wales, Australia collected between March 2020 and August 2021 were sequenced. Longitudinal samples from cases hospitalised due to SARS-CoV-2 infection (severe) were analysed and compared with cases that presented with SARS-CoV-2 symptoms but were not hospitalised (mild). SARS-CoV-2 genomic diversity profiles were also examined from daily sampling of culture experiments for three SARS-CoV-2 variants (Lineage A, B.1.351, and B.1.617.2) cultured in VeroE6 C1008 cells (n = 33).

**Results:** ISNVs were detected in 83% (19/23) of the mild cohort cases and 100% (16/16) of the severe cohort cases. SNP profiles remained relatively fixed over time, with an average of 1.66 SNPs gained or lost and an average of 4.2 and 5.9 low frequency variants per patient were detected in severe and mild infection, respectively. SgRNA was detected in 100% (25/25) of the mild genomes and 92% (24/26) of the severe genomes. Total sgRNA expressed across all genes in the mild cohort was significantly higher than that of the severe cohort. Significantly higher expression levels were detected in the spike and the nucleocapsid genes. There was significantly less sgRNA detected in the culture cohort than the clinical.

**Discussion and Conclusions:** The positions and frequencies of iSNVs in the severe and mild infection cohorts were dynamic overtime, highlighting the importance of continual monitoring, particularly during community outbreaks where multiple SARS-Cov-2 variants may co-circulate. SgRNA levels can vary across patients and the overall level of sgRNA reads compared to genomic RNA can be less than 1%. The relative contribution of sgRNA to the severity of illness warrants further investigation given the level of variation between genomes. Further monitoring of sgRNAs will improve the understanding of SARS-CoV-2 evolution and the effectiveness of therapeutic and public health containment measures during the pandemic.

## Introduction

As the ongoing Coronavirus disease 2019 (COVID-19) pandemic unfolds across the globe, several variants of concern (VOCs) of severe acute respiratory syndrome coronavirus 2 (SARS-CoV-2), the virus responsible for COVID-19 have emerged globally. Given the rapid worldwide spread, and continuing functional evolution of the virus, the real-time tracking of variants has become increasingly important (Wu et al., 2020, Zhou et al., 2020). SARS-CoV-2 genomes, approximately 30,000 bases in length are organised in a series of open reading frames (ORFs) consisting of four structural and 16 non-structural proteins (Arya et al., 2021). The 5’ end contains a leader sequence followed by the 5’ UTR and two large polyproteins (ORF1a and ORF1b) which encode all the non-structural proteins. These two ORFs are followed by the structural and accessory proteins, which include the spike protein (S), ORF3a, envelope protein (E), membrane protein (M), ORFs 6, 7a, 7b, 8, nucleocapsid (N), and ORF10 capped off by the 3’ UTR and poly-A tail (Nomburg et al., 2020, Naqvi et al., 2020). Following cytoplasmic entry into a host cell, the 1a and 1b large polyproteins are directly translated from genomic RNA (gRNA), while the remaining structural proteins are translated from sgRNA intermediaries (Sola et al., 2015, Song et al., 2019). The subgenomic RNA or sgRNA transcripts are produced through a complex mechanism involving discontinuous or ‘paused’ transcription, followed by an RNA-dependant RNA polymerase (RdRp) template switch during negative-strand RNA synthesis (Parker et al., 2021). The resulting nested set of negative sense RNAs serve as templates for the transcription of positive strands, forming mRNAs for translation of distinct proteins. sgRNAs contain a common leader sequence (65 to 90 nt) derived from the 5’ untranslated region, in addition to a transcription regulatory sequence (TRS) immediately adjacent to the 5’ open reading frame of the structural and accessory genes, responsible for the pausing of virus transcription during negative strand synthesis (Sola et al., 2015). The sgRNA of SARS-CoV-2 encode the structural proteins S, E M, and N, in addition to the several accessory proteins 3a, 6, 7a, 7b, 8, and 10 (Davidson et al., 2020, Kim et al., 2020).

Compared to DNA viruses, the replication of RNA viruses is typically associated with a high error rate due to the lack of sufficient proofreading activities during genome replication (Domingo and Holland, 1997). However, coronaviruses employ a highly conserved proofreading exoribonuclease encoded by non-structural protein 14 (nsp14) which enhances the fidelity of RNA synthesis (Graepel et al., 2017). Despite this mechanism, the mutation rate of SARS-CoV-2 is 1-2 mutations per month and is generally higher than DNA viruses.(Smith et al., 2014, Day et al., 2020, Nakagawa and Miyazawa, 2020). Additionally, coronaviruses have the propensity to recombine and generate extensive and diverse recombination products, particularly within the spike region of the genome (Wells et al., 2020).At an inter-host level, newly emerging viruses acquire adaptive mutations to enhance replication, modulate the host response, and facilitate effective transmission. However, the intra or within-host variability of RNA viruses is associated with the quasi-species concept, leading to multiple diverse circulating quasi-species of varying frequencies linked through mutation (Ramazzotti et al., 2020, Karamitros et al., 2020). The quasi-species collectively contribute functional characteristics at the population level, and in combination with the genetic profile of the host, can influence viral phenotype and adaptive capabilities (Stone et al., 2006). Since most of the immune escape and adaptive mechanisms of SARS-CoV-2 involve intra-cellular interactions, it is expected that SARS-CoV-2 evolves through intra-host selective pressure (Kumar et al., 2020), highlighting the capacity for the development of genetically different SARS-CoV-2 viruses within the same host.

Higher within-host diversity of viral RNA pathogens can be associated with increasing viral virulence and antigenic variability (Stone et al., 2006), exacerbated disease severity and clinical outcome, immune escape (Nowak et al., 1991), and drug resistance (Johnson et al., 2008). Given these effects, the importance and urgency of monitoring variants at the within-host level is paramount. Monitoring within-host diversity via the detection of intrahost single nucleotide variants (iSNVs) can inform genomic epidemiology (Lythgoe et al., 2021), and provide early indications of diagnostic PCR dropouts (Sapoval et al., 2020). The ability to predict mutations under positive selection, particularly functionally important and emerging mutations informs public health surveillance and the design of therapeutics (Wang et al., 2021, Popa et al., 2020, Tonkin-Hill et al., 2021).

Currently, variant analyses for SARS-CoV-2 focus primarily on mutations occurring at the consensus-level (single consensus sequence for each infected person), which represent the dominant variants within infected individuals (Lythgoe et al., 2021). However, genomic epidemiology of SARS-CoV-2 has revealed the capacity for viral mutations to emerge within an individual host (Lythgoe et al., 2021, Wang et al., 2021, Karamitros et al., 2020, Al Khatib et al., 2020), and so an understanding of the complete underlying within-host diversity at the population-level proves imperative.

This study aimed to investigate the consistency and timing of iSNV detection over the course of clinical and *in-vitro* SARS-CoV-2 infections using longitudinally collected specimens from the same patient. We examined iSNV profiles shared by SARS-CoV-2 lineages during an epidemiological characterised outbreak in Sydney, Australia. We investigated if these iSNVs were more frequently detected in severe illness, and if they develop over the time course of COVID-19 disease. We also measured changes in sgRNA to investigate if sgRNA is associated with increased genomic diversity during the course of individual infections.

## Materials and Methods

### Clinical specimens

Clinical specimens were collected 5 days prior to 23 days post onset of COVID-19 symptoms. If the date of symptom onset was unknown, the date of sample collection from the first positive specimen was considered the date of symptom onset. A total of 90 clinical specimens RT-qPCR positive for SARS-CoV-2 were included in the study. The majority of specimens were from the upper respiratory tract with nasopharyngeal swabs (n = 78), lower respiratory tract samples included bronchoalveolar lavages (n = 10) and sputum (n = 2) representing SARS-CoV-2 cases diagnosed in NSW, Australia between March 2020 and August 2021. The cohorts consisted of cases admitted to the intensive care unit +/-intubation (classified as severe disease) (n = 19 cases), and mild cases which recovered as outpatients (classified as mild disease) (n = 32). Nasopharyngeal swabs in Universal Transport Media (UTM) which were RT-qPCR negative for SARS-CoV-2 (n = 4) and collected in the study period were also included as negative controls. All specimens were de-identified and stored at -80°C. Following genome and variant level quality filtering the final cohorts consisted of 16 severe cases (n = 26 swabs) and 23 mild cases (n = 25 swabs).

### Ethics statement

Governance and human ethics approval for clinical metadata and use of specimens from cases positive for SARS-CoV-2 in New South Wales was obtained (2020/ETH02426 and 2020/ETH02282).

### Cultured isolates

Daily sampling was conducted to determine the fifty percent tissue culture infective dose (TCID_50_) of SARS-CoV-2 viral stocks from three lineages of SARS-CoV-2 (Lineage A – referred to as Wuhan, Beta – B.1.351, and Delta – B.1.617.2). Briefly, clinical samples confirmed to be SARS-CoV-2 positive by RT-qPCR (n = 3) were sequenced to determine the infecting SARS-CoV-2 lineage before being used for inoculation. CostarÒ 24-well clear tissue culture-treated multiple well plates (CorningÒ, Corning, NY, USA) were seeded at 40% confluence with Vero C1008 cells [Vero 76, clone E6, Vero E6] (ATCC® CRL-1586(tm)) in Dulbecco’s minimal essential medium (DMEM, Lonza Bioscience, Alpharetta, GA, USA), and supplemented with 9% foetal bovine serum (FBS, HyClone, Cytiva, Sydney, Australia). Culture media was changed within 12 hours and contained 1% FBS, and 1% antimicrobials including amphotericin B deoxycholate (25µg/mL), penicillin (10,000 U/mL), and streptomycin (10,000 µg/mL) (Lonza, Basel, Switzerland) to inhibit microbial overgrowth. The plates were inoculated with 200 μL of serially diluted virus stock (1×10^−2^ to 1×10^−6^) in triplicate. Cells were incubated at 37°C in 5% CO_2_ for 4 days (days 0 to 3), and were sealed with AeraSealä Film (Excel Scientific, Inc., Victorville, CA, USA) to minimise evaporation, spillage, and well-to-well cross-contamination. Visual inspection for the presence of cytopathic effect (CPE) was undertaken daily and 100 μL of supernatant was sampled from a single well to quantify viral replication every 24 hours. Mycoplasma testing was routinely conducted to exclude contamination of the culture media and cell line. The presence of CPE along with confirmation and quantification of SARS-CoV-2 viral load was performed by RNA extraction and RT-qPCR of culture supernatant daily (days 0 to 3) after viral load quantification RNA extracts were stored at -80°C or immediately used to prepare libraries for sequencing. All culture samples were identified via the following naming convention: <Lineage>-<day><dose> (i.e., A-D1-03; Lineage A, sampling day one, dilution 1×10^−3^). All SARS-CoV-2 culture was performed under level 3 biosafety conditions within a NSWHP physical containment level 4 laboratory (PC4) accredited facility.

### RNA extraction

Total RNA was isolated from mild clinical samples using the RNeasy Mini Kit (Qiagen) with minor modifications. A total volume of 200 μL of UTM /culture supernatant was added to 600 μL of RLT buffer and vortexed briefly. Next, 800 μL of 70% ethanol was added and mixed well by pipetting. The solution was then loaded on the RNeasy column in successive aliquots until the entire volume of the sample was extracted. RNA was eluted in 32 μL and stored at -80°C. Clinical samples from cases in severe (n = 48) and culture supernatants (n = 34) were extracted using the BioRobot EZ1 and EZ1 Virus Mini Kit v2.0 (Qiagen, Valencia, CA, USA) in PC4 facilities. An input volume of 100 μL was used and RNA was eluted into 60 μL as per the manufacturer’s instructions. The culture supernatants were extracted prior to removal of RNA from the PC4 facility.

### RT-qPCR of SARS-CoV-2

The SARS-CoV-2 viral RNA was detected and quantified using a previously described RT-qPCR targeting the N-gene (Rahman et al., 2020, Lam et al., 2021). Ten-fold serial dilutions (1 × 10^6^ to 1 × 10^2^ copies/µL) of the commercially available synthetic RNA control reference strain (Wuhan-1 strain, TWIST Biosciences) containing six non-overlapping fragments of the SARS-CoV-2 reference sequence (NCBI GenBank accession MN908947.3) was used to generate a standard curve and quantify SARS-CoV-2 viral load. The N-gene SARS-CoV-2 RT-qPCR was employed to determine the viral load of positive specimens sequenced as part of this study. The absence of SARS-CoV-2 in negative samples was confirmed by RT-qPCR.

### Virome enrichment, capture and sequencing

Viral enrichment of clinical extracts, *in-vitro* culture isolates, and the synthetic SARS-CoV-2 positive control spiked into negative culture supernatant was performed using the Illumina RNA Prep and Enrichment with the Respiratory Viral Oligo Panel (RVOP) v2 (Illumina, USA). This probe-based capture technique was selected as it was designed to generate near full length SARS-CoV-2 genomic sequences with even coverage in mutagenic regions. RNA denaturation, first and second strand cDNA synthesis, cDNA tagmentation, library clean-up and normalisation were performed according to the manufacturer’s instructions. Individual libraries were pooled in 3-plex reactions for probe hybridisation based on each samples SARS-CoV-2 viral load. The final probe hybridisation step was held overnight at 58°C. The enriched library was purified, and the concentration and fragment size were quantified using the Qubit™ 1x dsDNA High Sensitivity Assay (ThermoFisher Scientific, USA), and Agilent High Sensitivity D1000 ScreenTape assay on the Agilent 4200 Tapestation (Agilent, Germany), respectively. The libraries were sequenced using 2 × 74 bp runs on the Illumina MiniSeq™ or iSeq (Illumina, USA) and multiplexed with the aim of producing 2 × 10^6^ raw reads per library.

### Bioinformatic analysis and clustering

The raw sequence reads were subjected to an in-house quality control procedure prior to downstream analysis. The reads were demultiplexed and quality trimmed using Trimmomatic version 0.36 (minimum read quality score of 20, leading/trailing quality of 5). Reference mapping and consensus calling was performed using iVar version 1.2.1. Reads were mapped to the reference SARS-CoV-2 Wuhan-Hu-1 genome (NCBI GenBank accession: MN908947) and unmapped reads discarded. Mapping coverage and depth across the genome and structural and accessory genes was determined using MOSDEPTH version 0.2.9. Only genomes with >80% coverage over a 100x depth in all variant positions were included in further analyses. A consensus sequence was generated (map quality >20), and the 5’ (first 55 nt) and 3’ (last 100 nt) UTR regions were masked due to the known suboptimal sequencing quality of these regions. All genomes that passed filtering were submitted to NCBI GenBank (PRJNA633948). Phylogenetic Assignment of Named Global Outbreak LINeages (PANGOLIN) (https://github.com/hCoV-2019/pangolin) was used to infer SARS-CoV-2 lineages. In addition, isolates within lineages were assigned a genomic cluster denoting their position within NSW outbreak cases based on SNP distance from an epidemiological defined index case (Rockett et al., 2020).

### Variant filtering and analysis

The frequency and positions of variants (SNPs and iSNVs) in all samples were determined using Varscan version 2.3.9. SNPs were defined as mutations with a read frequency of ≥ 0.9. Variants with a read frequency between 0.05 and 0.9 were defined as low frequency variants. Variants occurring in the TWIST control were highlighted as potential artefacts (MN908947 genome reference positions 5765, 5766, 1107, 11082, 12413, 12926, 23652, and 26433), and those associated with mis-mapping at the ends of insertion or deletion events in B.1.351 and B.1.617.2 (11288, 22029, 22034, 22287, 23598, 23607, 23609, 23616, 28248 and 28249) were identified and excluded from downstream analyses. Changes in the iSNV profiles were determined between longitudinal sampling from individual cases, cases from the same household (known transmission events), within severe and mild cohorts, across lineages, and genomic clusters.

### Detection of SARS-CoV-2 subgenomic RNAs

SgRNA analysis was conducted using Periscope (Parker et al. 2021). Briefly, Periscope distinguished sgRNA reads based on the 5’ leader sequences being directly upstream from each genes transcription. The sgRNA counts were then normalised into a measure termed sgRPTL, by dividing the sgRNA reads by the mean depth of the gene of interest and multiplying by 1000.

### Phylogenetic analysis

Consensus SARS-CoV-2 genomes were processed using the Nextstrain Conda environment. Augur (bioinformatics tool) v13.0.0 (https://github.com/nextstrain/augur), and Auspice (open-source visualisation tool) v2.30.0 (https://github.com/nextstrain/auspice) were employed for analysis and visualisation (Hadfield et al., 2018). As a comparison, a representative global subset of SARS-CoV-2 genomes curated by Nextstrain between September 2019 and August 2021 was included in our phylogenetic analysis.

## Statistical analysis

Statistical significance (*p* ≤ 0.05) was determined using the Mann-Whitney test for difference between means on variables which contained at least five data points (iSNV/SNP counts and read frequencies, sgRNA counts and read frequencies).

## Results

### Phylogenetic analysis

In total, 84 high-quality SARS-CoV-2 genomes collected between December 2019 to August 2021 were added to a sub-sample of global SARS-CoV-2 genomes (n = 1000). These global strains were collected using a sampling strategy to obtain a representative dataset across all regions (Asia, Oceania, Africa, Europe, North America and South America). The completed build was used to generate a .json file containing 1,163 viral genomes. This file was then run in Auspice, the program to render Nextstrain visualisations, creating a radial divergence phylogeny (Figure 1).

**Figure 1:**
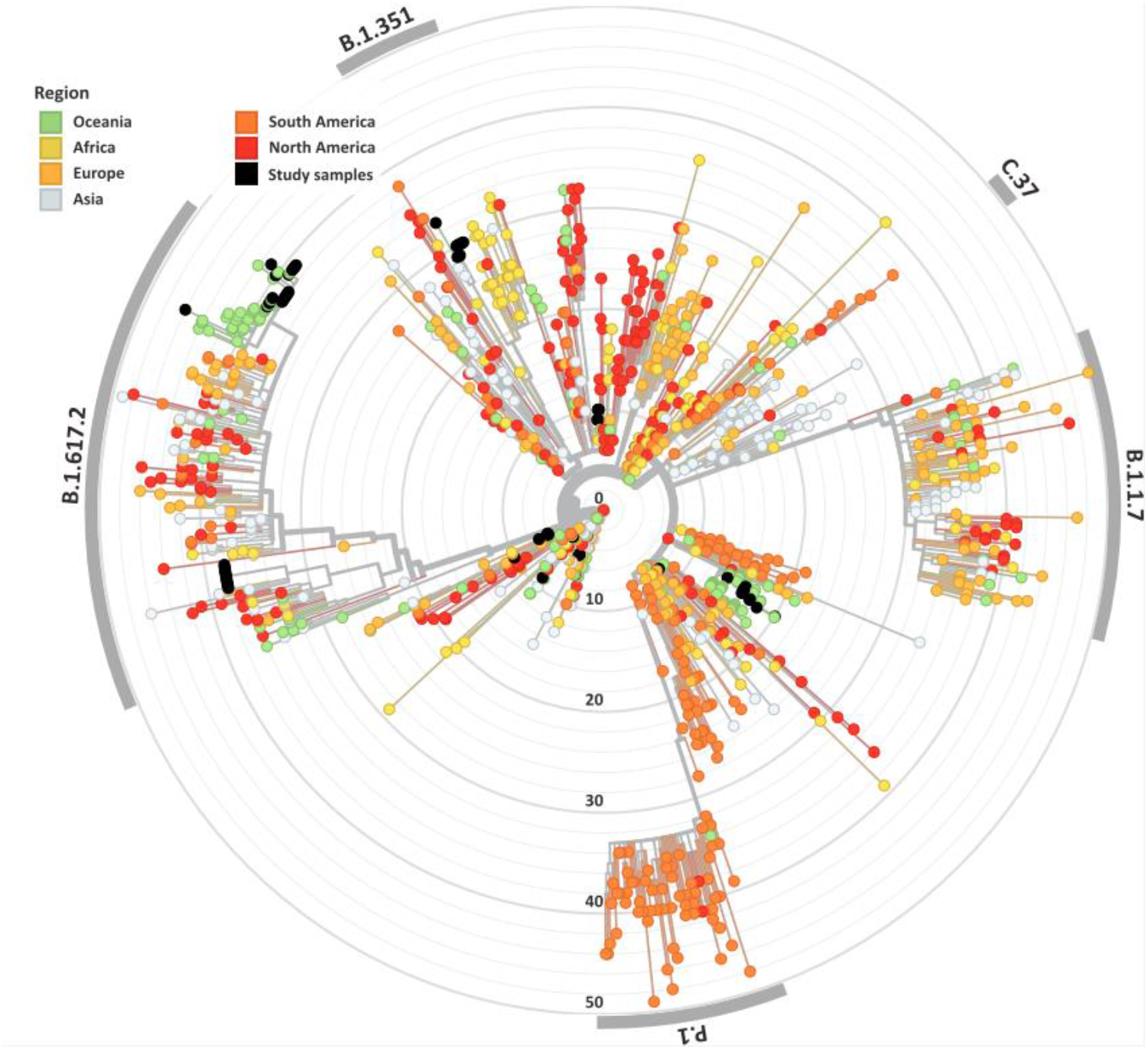
Genomic and epidemiological diversity of clinical and culture SARS-CoV-2 sequences. The SARS-CoV-2 phylogeny was constructed in a local version of Nextstrain (auspice.us) and shows the evolutionary relationship of SARS-CoV-2 genomes sequenced within this study (black dots) and a subsample of publicly available SARS CoV-2 genomes collected globally (n = 1,079). The tree branch lengths represent divergence, with sequences rooted relative to early samples collected in Wuhan, China (Wuhan-Hu-1/2019).

### Cohort sequencing results

After genome and variant-level filtering and quality control, 25 SARS-CoV-2 consensus genomes were recovered from the mild (n = 23 cases, average sample per case = 1.1) and 26 from the severe (n = 16 cases, average sample per case = 1.6) cohorts (Table 1). There were no significant differences between age and sex across the mild and severe cohorts, however, a significant difference between the age ranges (*p* = 0.16) was noted (Table 1). Thirty-three SARS-CoV-2 consensus genomes were sequenced from 34 culture specimens of varying sample dilutions and time intervals (one genome did not pass quality filtering and was excluded from the analyses). High depth genomes were produced across all cohorts and the median depth achieved was not significantly different. The median depth for the severe cohort was 2,021x, mild 928x, Lineage A 2,964x, Beta VOC 3,408x, and Delta VOC 2,529x (Supplementary Figure S1b).

**Table 1:** Demographics of SARS-CoV-2 positive cases within the severe and mild cohorts.

| **Cohort** | **Total Cases** | **Total Samples** | **Lineages (n samples)** | **Gender M : F** | **Age Median Range** |
| --- | --- | --- | --- | --- | --- |
| **Severe disease** | 16 | 26 | B.1 (7), B.6 (4), B.1.617.2 (13), A.2.2 (1), A (1) | 8M : 8F | Median = 65; Range 27-94 |
| **Mild** | 23 | 25 | D.2 (11), B.1 (6), B.1.617.2 (7), B.1.1 (1). | 6M : 15F | Median = 56; Range 12-71 |
| **Total** | 39 | 51 | A (1), A.2.2 (1), B.1 (13), B.1.1 (1), B.1.617.2 (20), B.6 (4), | 14M : 23F | Median = 58; Range 12- 94 |

## SARS-CoV-2 viral load

A range of SARS-CoV-2 viral loads were detected in each cohort (Supplementary Figure 1a). The median severe viral load was 516,643 copies (range: 151,246,026 to 2,512 copies), and the median mild viral load was 457,284 copies (range: 95,727,865 to 668.7 copies). Within the culture cohorts, lineage A median viral load was 1,408,340 copies (range: 18,976,383 to 29.4 copies), Beta median viral load was 560,453.7 copies (range: 9,026,044 to 130.2 copies), and Delta median viral load 300,232.9 copies (range: 3,107,421 to 424.5 copies) (Supplementary Figure S1a). There was no significant difference between the viral loads across cohorts.

### SARS-CoV-2 Pango lineages and epi-clusters

A wide variety of pango lineages were defined across the clinical cohorts (Table 1). The majority of cases were designated to lineage B.1.617.2 or Delta VOC (n=20, cases; P601, P603, P608, P611, P612, P614, P615, P622, P624, P626, P629, P604, P606, P618, P620, P621, PG27, P628), followed by B.1 (n=13 cases P0332, P0570, P0495, P0676, P1384, P1434, and P1494), D.2 genomes (n=11, cases; P0340, P0341, P0417, P0642, P0858, P1149, P2099, P2152, P1498 and P1551) and B.6 (n=4, case P0105). Single genomes from lineage A.2.2 (case P1727) lineage A genome (case P1811) and lineage B.1.1 (P1020) (Figure 1, Table 1). Within some lineages, genomes were designated distinct epi-clusters based on SNP distances and known epidemiological links from public health investigations. Within Lineage B.1.617.2 there was one cluster (NSW 130, n = 20), B.1 had 3 clusters (NSW 17.5, n= 3; NSW 9, n = 7, and singletons n = 2), B.6 had 1 cluster (NSW 3.1, n = 4), and D.2 had 2 clusters (NSW 33.1, n = 6; NSW 33, n = 7). Within the epi-clusters known transmission events between members within a household were also captured; group 1 lineage B.1 (n = 3 cases, 3 samples), group 2 lineage D.2 (n = 2 cases, 2 samples), group 3 lineage B.1.617.2 (n = 2 cases, 3 samples), group 4 lineage B.1.617.2 (n = 2 cases, 2 samples), and group 5 lineage B.1.617.2 (n = 2 cases, 2 samples).

### Frequency of iSNVs across cohorts

Low frequency iSNVs were detected in 100% (16/16) of the severe (Figure 2a), 83% (19/23) of the mild (Supplementary Figure S2), and 100% (33/33) of the culture samples (Figure 2b). Longitudinal samples collected from the same patient were collected over a mean of 6.36 days (range: 0 to 11) post-symptom onset compared to one to three days after inoculation in cultured specimens. Overall, there were 91 iSNVs detected in the severe cohort (median number of iSNVs per sample per case = 3, median frequency = 0.106), and 129 iSNVs detected in the mild cohort (median number of iSNVs per sample per case = 4, median frequency = 0.085) (Supplementary Figure S2). The frequency of iSNVs per SARS-CoV-2 gene between severe and mild cohorts was significantly different only for the S gene (*p* = 0.0209) (Figure 3). Of the culture specimens there were 60 iSNVs (median frequency per specimen= 0.373) for the lineage A cultures, the Beta culture contained 39 iSNVs (median frequency = 0.345), and the Delta culture contained 39 iSNVs (median frequency = 0.214) (Figure 2b).

**Figure 2:**
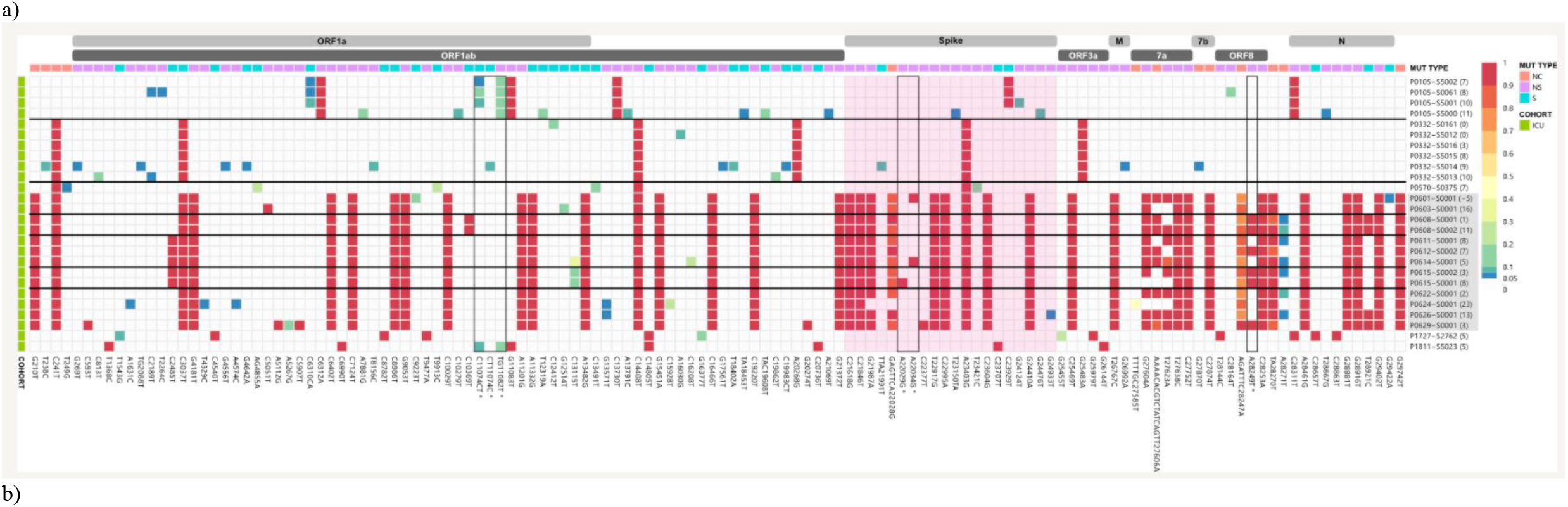
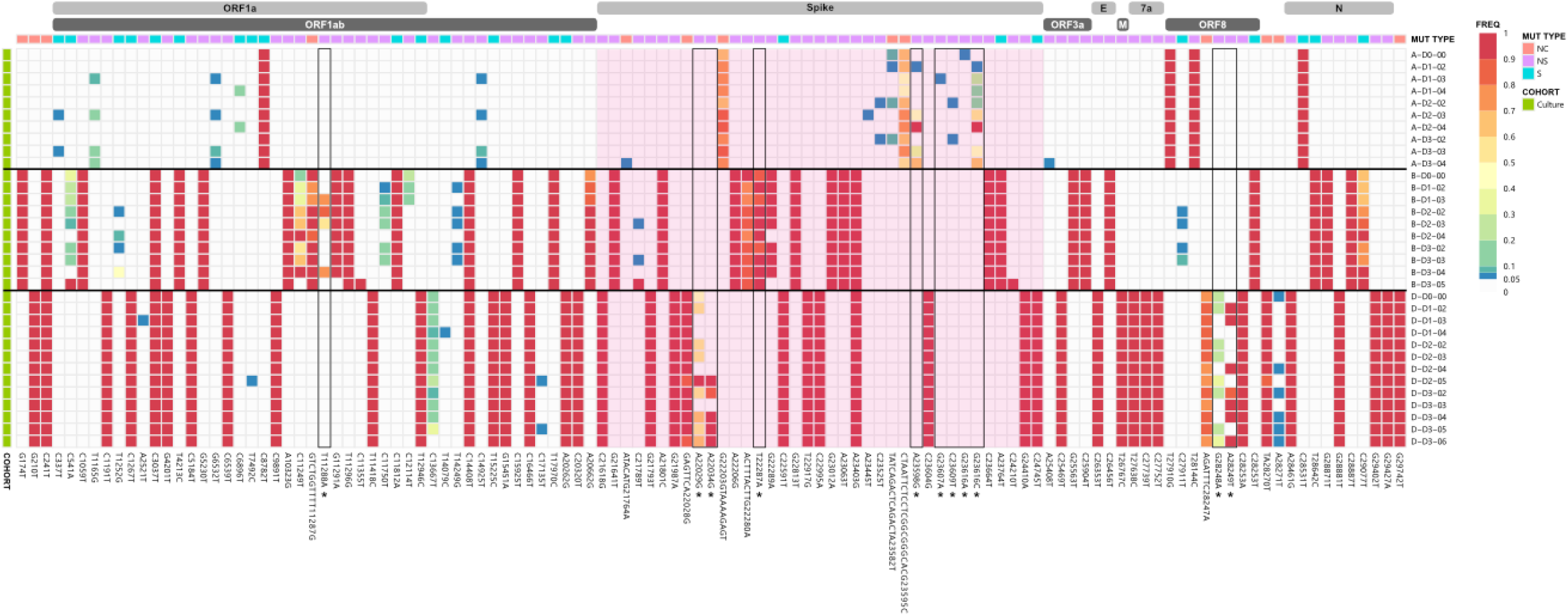
iSNV and SNP frequency and the corresponding synonymous and non-synonymous mutations across the SAR-CoV-2 genomes for the a) severe and b) culture cohorts. A frequency of ≥ 0.9 was considered a SNP (red), frequencies below 0.05 were not included in the analysis. Identified problematic sites are outlined in black and denoted with an * on the x axis. The spike gene is highlighted in pink. SARS-CoV-2 Delta lineages are highlighted in grey. Bracketed numbers indicate the date of sample collection post-symptom onset.

**Figure 3:**
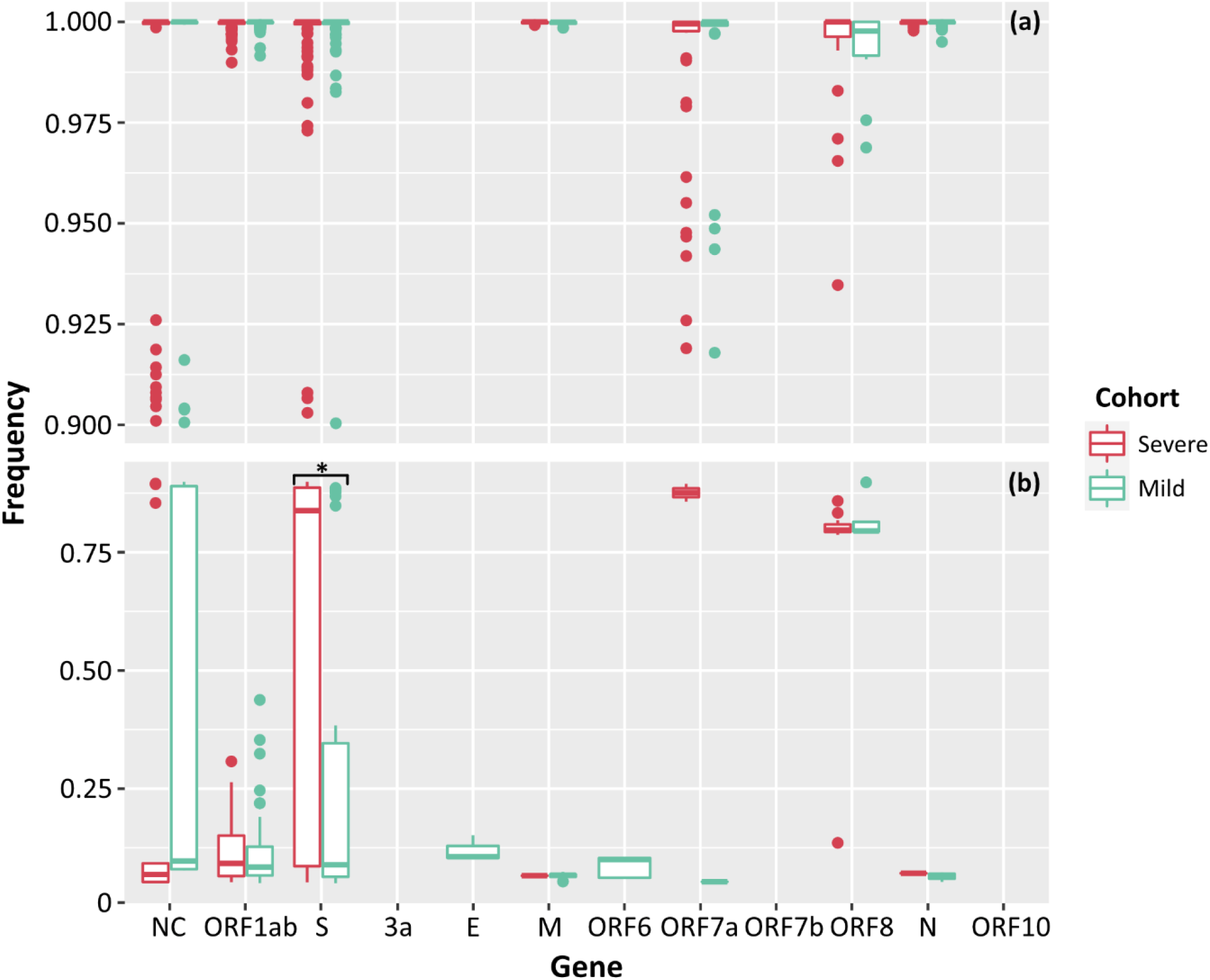
Frequencies of SNPs (a) and iSNVs (b) by SARS-CoV-2 gene for severe (red), and mild (green) cohorts. Frequencies ≥ 0.9 were considered SNPs. Problematic sites are not included. NC signifies non-coding region of the genome. Statistical significance (*p* ≤ 0.05) is denoted by (*). The frequency of iSNVs in the spike gene of SARS-CoV-2 were significantly different between the severe and mild cohorts.

There was no significant difference between the median numbers of iSNVs per case when comparing the severe and mild cohorts. However, a significantly higher median read frequency of iSNVs between the severe and mild cohorts (*p* = 0.023) was observed. There was also a significant difference between the mean frequency of iSNVs between the severe and culture cohorts (*p* = 0.016), and the mild and culture cohorts (*p* = 0.00001). There was also a significantly higher number of iSNV in the lineage A culture compared to the Delta variant culture (*p* = 0.00001). The highest number of SNPs and iSNVs/per SARS-CoV-2 genome in cases for all cohorts were found in ORF1ab, S, ORF8 and N genes. Differences between the counts of iSNVs between severe and mild cohorts were significant only at the ORF1ab gene (*p* = 0.0357) (Figure 4). Across all cohorts, the number of non-synonymous SNP and iSNV mutations were greater than synonymous, indicating positive selection. The average non-synonymous/synonymous mutation ratio (Ka/Ks) per genome per cohort for iSNVs and SNPs was 2.30 and 3.33 in culture, 1.47 and 5.29 in severe cases, and 1.65 [1.63 excluding outlier] and 2.84 in the mild cohort. The SNP ka/ks ratio was highest across all cohorts when compared to the iSNV ka/ks ratio.

**Figure 4:**
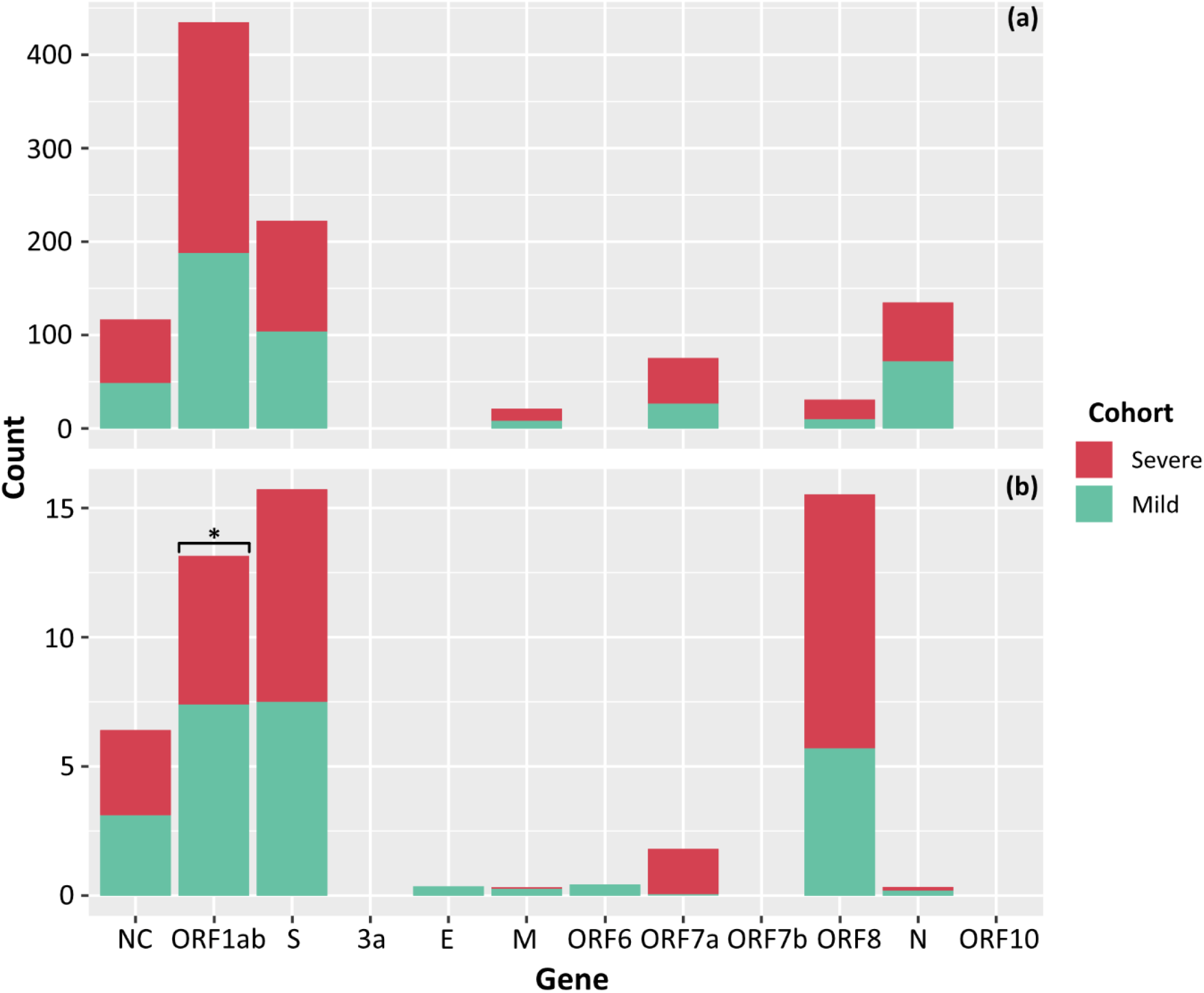
Counts of SNPs (a) and iSNVs (b) by SARS-CoV-2 gene for severe (red), and mild (green) cohorts. Frequencies ≥ 0.9 were considered SNPs. Problematic sites are not included. Statistical significance (*p* ≤0.05) is denoted by (*) and NC signifies non-coding region of the genome. The number of iSNVs in the ORF1ab gene of SARS-CoV-2 were significantly different between the severe and mild cohorts.

**Figure 6.**
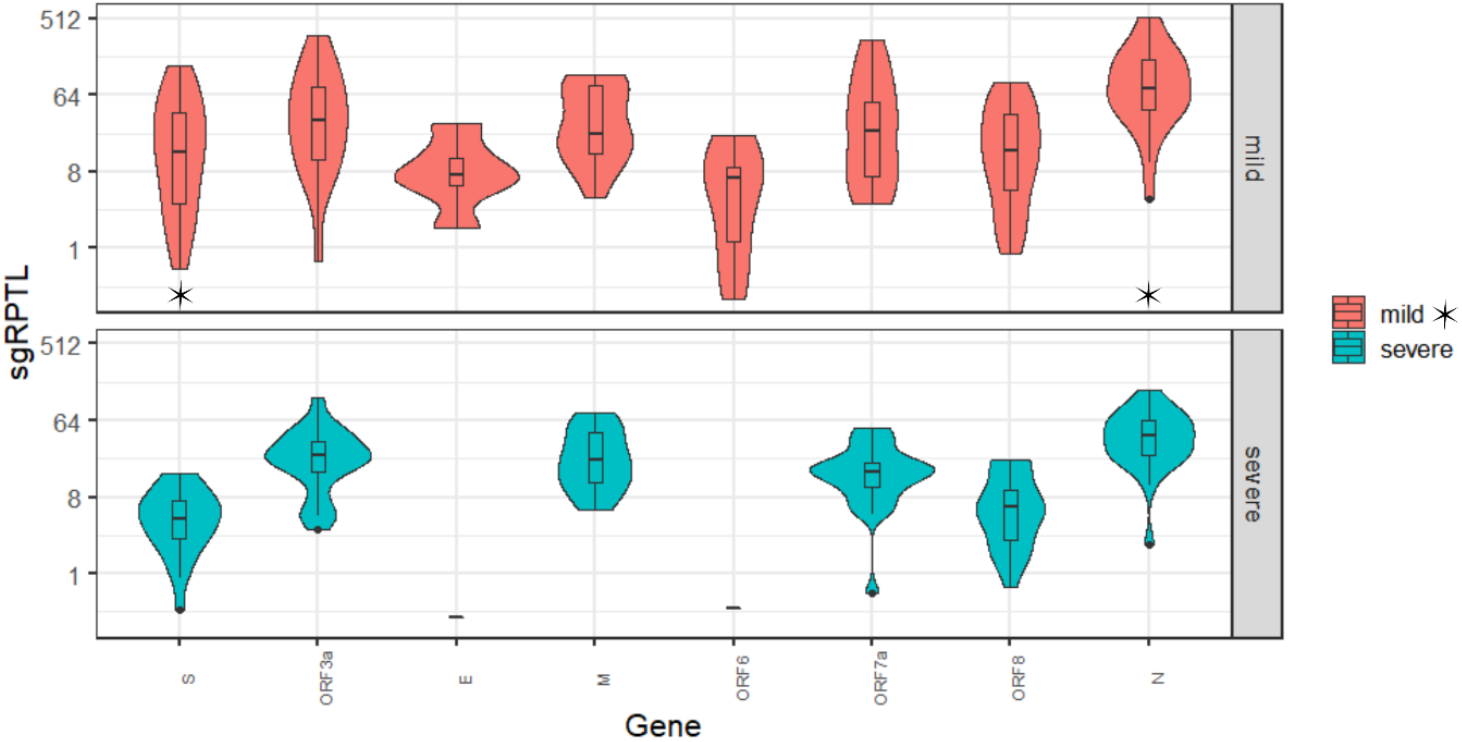
Distribution of sgRPTL counts across all genes sgRNA was detected in in the mild (red) and severe (green) clinical cohorts. Statistical significance (*p* ≤0.05) is denoted by (*). S sgRNA detected in n = 24/26 severe and n = 21/25 mild genomes, ORF3a in 24/26 severe genomes and 17/25 mild genomes, E in 1/26 severe genomes and 6/25 mild genomes, M in 20/26 severe genomes and 15/25 mild genomes, ORF6 in 1/26 severe genomes and 11/25 mild genomes, ORF7a in 24/26 severe genomes and 18/25 mild genomes, ORF8 in 22/26 severe genomes and 20/25 mild genomes and N in 25/26 severe genomes and 24/25 mild genomes.

### Clinical Longitudinal samples measuring Intra-host diversity

Our cohort contained four severe cases with longitudinal samples (P0105, P0332, P608 and P615), with an average of 3.25 samples per case. The SNP profile of each patient remained relatively fixed over time, with an average of 1.66 SNPs gained or lost (range 1 - 2) compared to the initial genome collected for each case. There was a general trend observed with an increase in the number of iSNVs over the course of the earlier lineage infections. For severe case P0105, a single iSNV (genome position 6310 nt, read frequency < 0.12) remained consistent across three longitudinal samples which were seven to ten days post symptom onset. However, this iSNV was lost at 11 days post symptom onset. Despite the loss of the iSNV on day 11 (position 6310 nt), this sample contained iSNVs (frequencies < 0.3) at eight new locations within the genome; five in ORF1ab, two in spike, and one in nucleocapsid. Severe case P0332 had no consistent iSNVs throughout their infection, although an average of three iSNVs (range: 0 - 13) were detected per sample. Days nine and ten post symptom onset (last two sampling points) contained the greatest diversity of iSNVs, with iSNVs present at 12 positions on day nine (ORF1ab, n = 9; S, n = 1; M, n = 1; and NC, n = 1) and three positions on day ten (ORF1ab n = 3). In B.1.617.2, there were fewer conserved iSNVs and iSNVs tended to be lost rather than gained. For case P608 one iSNV and one low frequency deletion event was retained from one day to 11 days post symptom onset (NC, n = 1; ORF8, n = 1). In addition, one iSNV present at one day post symptom onset converted to a SNP at 11 days post symptom onset (NC, n = 1) and one high frequency deletion event at day one converted to a low frequency deletion event by day 11 (S, n=1). The final longitudinal sample, severe case P0615, retained one iSNV and two low frequency deletion events from three days post symptom onset to eight days post symptom onset (ORF1ab, n = 1; S n = 1; NC, n = 1) with one iSNV lost at day 8 (NC, n = 1).

Our cohort also contained five epi-linked family groups. There were no shared iSNVs between cases in groups 1 and 2 (Lineages B.1 and D.2). Group’s three to five were all in lineage B.1.617, and of those groups, groups four and five had no shared iSNVs between cases. In group three there was one iSNV shared amongst all cases and samples (ORF1ab, n = 1), and one iSNV that was present in one case and gained later in the course of infection in the other case (NC, n = 1). However, in all three groups there were two shared deletion events (S, n = 1; NC, n = 1).

### Culture intra-experiment genomic diversity

In culture, SNPs remained relatively stable over sampling time-points and across lineages and dilutions with zero SNPs in lineage A, 15 SNPs and four high frequency deletion events in lineage Beta, and one SNP and two high frequency deletion events in lineage Delta were lost or gained over the three-day experiment. There was overall greater diversity of iSNVs occurring within the Beta and Delta VOCs when compared to lineage A (60 and 39 compared to 20 with median frequencies of 0.288 and 0.153 compared to 0.0663). There was a significant difference between the median frequencies of lineage A compared to Beta (*p* = 0.00001) and Delta (*p* = 0.00214) but no significant difference between Beta and Delta. Large indels were noted in the lineage A spike gene (22203 to 22213 nt, and 23595 to 23585 nt) and remained at relatively high frequency (> 0.4) over time and dilutions. Lineage-specific deletions in Beta (position 22270 to 22280 nt) and Delta (position 28240 to 28247 nt) cultures were maintained over the time course of the experiment but remained at frequencies less than 0.9. Interestingly, iSNVs that occurred *in-vitro* were not present at baseline (inoculum sample) 88% (8/9), 75% (6/8), and 71% (5/7) of the time in lineage A, Beta, and Delta variants, respectively. These iSNVs were often 96% (45/47) below a frequency of 0.3.

### Within-host genomic diversity between lineages and epidemiologically defined transmission

The iSNVs were higher within the B.1, n = 12 genomes (39 positions along the genome, with an average of 1.025 iSNVs/position), and D.2 lineages, n = 11 genomes (45 positions along the genome, with an average of 1.666 iSNVs/position), and B.1.617.2 n = 13 genomes (29 positions along the genome, with an average of 1.714 iSNVs/position). Lineages with <5 genomes were excluded. Within those lineages iSNVs were most commonly shared between genomic clusters in the D.2 lineage and were only shared once between a singleton and cluster 9 lineage B.1. iSNVs were shared between lineages B.1 and D.2 at eight positions (ORF1ab – 269 nt, 3,761 nt, 5,372 nt, 6,604 nt, 11,511 nt; ORF3a – 25,408 nt; M – 26,545 nt; and NC – 27,870 nt). There was one occasion where a SNP in lineage B.6 was a 0.05 frequency iSNV in another (D.2) (Figure 2).

### Subgenomic RNA

Subgenomic RNA was present in almost all genomes from severe and mild cohorts (24/26, 25/25), although at low levels, median 1.7% of the read depth compared to gRNA (range; 0.02% - 52% of depth/average gene depth). The N-gene was the highest sgRNA transcript detected in the severe cohort measured by median sgRPTL (severe 41.916; mild 76.25) followed by the M gene (22.309; 22.549), ORF3 (24.873; 32.389), ORF7a (15.867; 24.259), ORF8 (6.227; 14.386), and S (4.353; 13.422). Only a small number of genomes expressed sgRNA for ORF1ab (0.849 n=1; 2.87 n = 2), ORF6 (0.386 n = 1; 6.623 n = 11) and E (0.311 n = 1; 7.335 n = 6), no sgRNA was detected for ORF10. There was a significant difference (*p* = 0.00804) between the sgRNA across all genes of the severe and mild cohorts – comparing individual genes where both had a sample size > 5, there were no significant differences except in the N (*p* = 0.0114) and S (*p* = 0.0128) genes (Figure 5). Although there were also some interesting trends in sgRNA between lineages, the sample sizes per lineage were insufficient to determine significance (Supplementary Table S1). Within the culture cohort, sgRNA was also present in the majority of genomes (30/33) at low levels with a median of 0.8% of the total reads compared to gRNA (range 0.02% - 10.4% depth/ average gene depth). Overall, sgRNA expression was significantly less than in both the severe (*p* = 0.0002) and mild (*p* = 0.0001) cohorts (Supplementary Figure S3).

## Discussion

### Intra-host variation

Since the emergence of SARS-CoV-2 in 2019 the virus has been continually acquiring polymorphisms. In some cases, these changes have resulted in VOCs that pose a greater threat to public health, in the form of increased transmissibility and evasion of current prevention strategies (Wu et al., 2020). The evolutionary rate of SARS-CoV-2 has been estimated at 1.1 × 10^−3^ substitutions/site/year (Smith et al., 2014, Day et al., 2020), however, some variants, such as Alpha (Lineage B.1.1.7) and Delta (Lineage B.1.617.2) have developed a higher percentage of nucleotide changes (Duchene et al., 2020, Rambaut, 2020). Low frequency iSNVs within the viral population of an affected host are thought to be a contributing factor to the emergence of mutations. These iSNVs have been described across the entirety of the genome and can affect both non-protein coding and protein coding genes (Karamitros et al., 2020, Lythgoe et al., 2021, Armero et al., 2021). In this study we documented that the majority of iSNVs detected in the clinical samples were present at low frequencies (average: 0.0795) and were not present in longitudinal samples. To accurately quantify iSNVs we sampled longitudinal samples collected from 6 cases and 3 known transmission events. Cases P0105 and P0332 had severe infections and had variants present on the day of symptom onset at reference position 12,412 nt (synonymous, gene ORF1a I4049I) that reverted, but subsequent samples 10 days post symptom onset detected iSNVs at position 19,862 nt (non-synonymous, gene ORF1b A2132V). Where consistent iSNVs were present across longitudinal samples, they were at low frequencies that were subsequently lost. For example, patient P0105 at position 6,310 nt (insertion, gene ORF1a 2015), iSNVs were present at seven, eight-, and 10-days post symptom onset, but were no longer detectable at 11 days post symptom onset (final sampling). This appears to be consistent with previous studies which demonstrate transmission bottlenecks, where the majority of iSNVs are eventually lost and not transmitted onto new individuals (Lythgoe et al., 2021, Valesano et al., 2021). However, there may be implications for transmission and potential emergence of VOCs if iSNVs evolve into SNPs. Our findings indicate that the number of iSNVs tends to increase the longer the infection progresses, particularly in the ORF1ab, S, and N genes. It is therefore possible that the longer an individual remains infectious, the higher the likelihood of the accumulation and transmission of functional iSNVs.

It is important to note that the presence and/or absence of iSNVs and their distribution across the genome changes from patient to patient and within cases over time. There are some consistently polymorphic positions-for example position 23, 929 nt in lineage B.6 is a consensus level change but is commonly an iSNV in the D.2 lineage. Lineage D.2 and B.1 in particular, had high levels of iSNVs respectively and high levels of shared iSNV positions in the genome. All samples from lineages D.2 and B.1 were collected from cases of local transmission whereas infection in cases associated with lineages A and B.6 were acquired overseas. This supports the recent report by Armero et al. 2020 of high levels of carryover iSNV diversity in the ORF1ab, S, and N genes within an outbreak in Victoria, Australia.

In contrast when SARS-CoV-2 was grown within *in-vitro* culture systems and sampled at consistent timepoints, the location of iSNVs was conserved and present at a significantly higher read frequency than within the clinical cohorts. Within the Beta culture, multiple iSNVs in roughly 50% of the reads at inoculation became a high frequency iSNV (in > 80% of reads) or a SNP (≥ 90% of reads) by day three. We also documented instances where iSNVs were present at low frequencies at inoculation (baseline sampling) and remained at a low frequency across the study period, except for one dilution in which a SNP developed at day two and persisted. The Delta culture contained an iSNV that was present at inoculation, lost in all dilutions at day one, returning in one dilution on day two and then became present in all but one dilution by day three. Interestingly, there were no iSNVs within the S gene of the Delta lineage. Therefore, the presence of an iSNV early on in infection does not ensure that the variant will remain throughout the patient’s course of infection, consistent with what has been observed in prior work (Armero et al., 2021, Lythgoe et al., 2021). It is also evident that a lack of iSNVs early on in infection does not indicate that a mixed population will not arise at some point during the infection course. This observation has implications for interpreting relationships between genomes when iSNVs are used to trace chains of transmission. There was also an interesting change in the representation of deletion events at the sub-consensus level between culture lineages. Within lineage A there were two low frequency deletion events within the S gene that overlapped, positions 23,583 to 23,598 nt, and 23,596 to 23,617 nt where the first deletion was at a considerably lower frequency than the second. This is indicative of viral evolution towards the most advantageous. These polymorphisms are concentrated near the furin cleavage site and occurred predominately when SARS-CoV-2 is grown in VeroE6 cells. This cell line lacks key proteinases that enable more efficient viral entry and fusion (Chaudhry et al., 2020).

While most mutations are purged or have no effect on the fitness of the virus, some may be selected for and alter transmissibility, infectivity, or pathogenicity (Plante et al., 2021). In both the clinical and culture cohorts over 50% of the iSNVs resulted in a non-synonymous change, and between 21 to 37% iSNVs indicated a synonymous change. In all instances, positive selection pressure was observed. Coronavirus mutations in the functionally important spike protein have the potential to affect virus infectivity, pathogenicity, and susceptibility to neutralizing antibodies (Harvey et al., 2021). The spike gene encompasses positions 21,563 to 25,384 nt and iSNVs were seen within this range in all cohorts at the second highest frequency, with only ORF1ab being higher. This is consistent with studies that have observed positive pressure on protein coding genes, especially those associated with surface glycoproteins (Lo Presti et al., 2020).

### Subgenomic RNA variation

We uncovered low levels of sgRNA expression across all three cohorts, representing, on average less than 2% of the read depth of gRNA. However, the relative abundance of the eight sgRNA transcripts was similar to other investigations, where nucleocapsid sgRNA transcripts were the most abundant and ORF10 sgRNA was not detected (Alexandersen et al., 2020, Parker et al., 2021). Interestingly the pattern of sgRNA detection was similar across cohorts and we did not detect a higher abundance of sgRNA in cultured isolates, as previously reported (Nomburg et al., 2020). Instead, we found a significantly higher level of sgRNA in cases with mild symptoms. This is an interesting result as it has been reported that sgRNA transcripts are reduced in asymptomatic cases of COVID-19 (Wong et al., 2021). However, this may be a result of the lack of intervention in mild cases compared to interventions which would have been received by hospitalised severe disease cases. This similar pattern of expression also remained unchanged between viral lineages (D.2, B.1 and B.1.617.2). The presence of sgRNA transcripts in the E and N regions can be considered markers for increased replication (Zollo et al., 2021). However, it was postulated (Alexandersen et al., 2020) that levels of sgRNA may not be a reliable indicator of disease progression. Our data supports this assumption with levels of sgRNA in genes of importance, such as S and N remaining relatively even across the longitudinal clinical samples. Further to this concept, the median levels of sgRNA detected in the severe cohort were less than that detected in the mild cohort in all genes except M. This is inverse to the assumption that cases in the severe cohort are generally considered to have high levels of sgRNA. It is possible that the production of sgRNAs is more relevant for transmission, cell entry and less important in viral propagation. In addition, trinucleotide mutations have been identified in some lineages generating novel TRS which increases expression of sgRNA transcripts. This can be seen in the B.1 and D.2 lineages which expressed the highest levels of sub genomic transcripts for the nucleocapsid encoded by a GGG > AAC mutation (nt 28881 – 28883). It is still unknown how these new transcripts will impact pathology, but it is hypothesized that it could lead to diversification and adaptation to the host (Long, 2021).

We have established significant differences between severe and mild disease cohorts and genes as well as distinct and consistent patterns of sgRNA. Our findings are also consistent with relative abundances of sgRNA described at spike and nucleocapsid genes (Alexandersen et al., 2020). However, this study was limited in the available sample size, which was further complicated by low viral levels in later longitudinal samples. Strict lockdown procedures and border closures also greatly reduced or eliminated the proliferation of SARS-CoV-2 lineages, leading to low numbers of representative genomes per lineage. Further investigations with larger time frames and more longitudinal samples will be required to fully understand the behaviour and contribution of iSNVs to disease and transmission.

## Conclusion

We demonstrate that iSNVs in SARS-CoV-2 genomes can accumulate over the course of COVID-19 infection and were predominately sporadic across cases with severe or mild disease. There were lineage-specific hot spots associated with persistent and low level iSNVs within diverse samples. sgRNA expression appears relatively consistent across both severe infections and mild infections with the exception significantly higher expression of sgRNA spike and nucleocapsid transcripts in the mild disease cohort. The levels of sgRNA were, on average, less than two percent of the total reads for any gene in any clinical sample, indicating that SARS-CoV-2 sgRNA may not be a major contributor to the severity of clinical presentations of COVID-19. The ongoing surveillance and monitoring of subpopulations and iSNVs within lineages over time can improve our understanding of the underlying SARS-CoV-2 host adaptation. In addition, monitoring of sgRNA levels, especially associated with severity of disease may be important in understanding their impact on the spread of COVID-19.

## Data Availability

Fastq files have been deposited in the National Center for Biotechnology Information Short Read Archive BioProject PRJNA633948 for all 84 genomes produced in this study.

## Acknowledgements

The authors thank the staff from the Virology Laboratory and Microbial Genomics Reference Laboratory at the Institute of Clinical Pathology and Medical Research, NSW Health Pathology for their logistical support, and the Sydney Informatics Hub, University of Sydney Core Research Facility at the University of Sydney for providing research computing services. The authors also thank partner laboratories within the NSW Health Pathology network for their participation in the COVID-19 genomic surveillance, and GISAID team for their efforts in collating global data on the SARS-CoV-2 pandemic.

## Supplementary Material

**Supplementary Figure S1:**
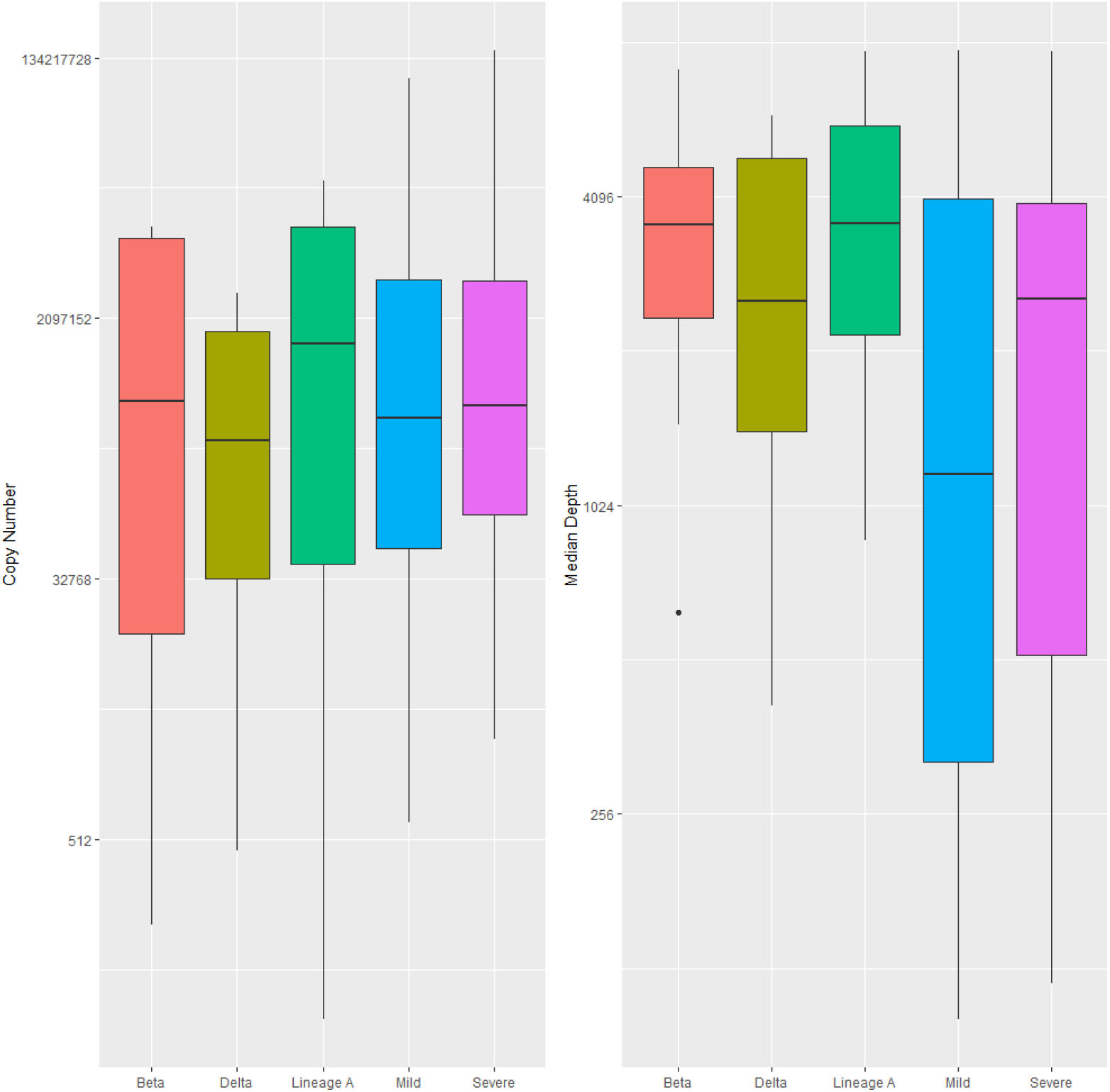
Boxplots depicting the copy number a) and coverage b) for all cohorts. The bold line indicates the median, the interquartile range (25th to 75th percentile) is represented by the white shading, the whiskers represent the minimum and maximum values, and the outliers are shown 759 by the black circles.

**Supplementary Table S1:** Median sgRPTL by gene for lineages with a sample size greater than five.

| Lineage | Gene |  |  |  |  |  |  |  |  |
| --- | --- | --- | --- | --- | --- | --- | --- | --- | --- |
|  | ORF1ab | S | ORF3a | M | ORF7a | ORF8 | N | ORF6 | E |
| **B.1** | 5.474 | 12.452 | 95.553 | 22.549 | 42.748 | 13.674 | 60.246 | 0.397 | 0 |
| **B.1.617.2** | 0 | 2.987 | 13.617 | 15.383 | 11.957 | 5.262 | 39.946 | 0.386 | 0.311 |
| **B.6** | 0 | 7.848 | 32.987 | 33.102 | 20.912 | 6.126 | 0 | 0 | 0 |
| **D.2** | 0.273 | 38.501 | 56.811 | 86.419 | 84.859 | 39.981 | 139.286 | 7.173 | 7.335 |

**Supplementary Figure S2:**
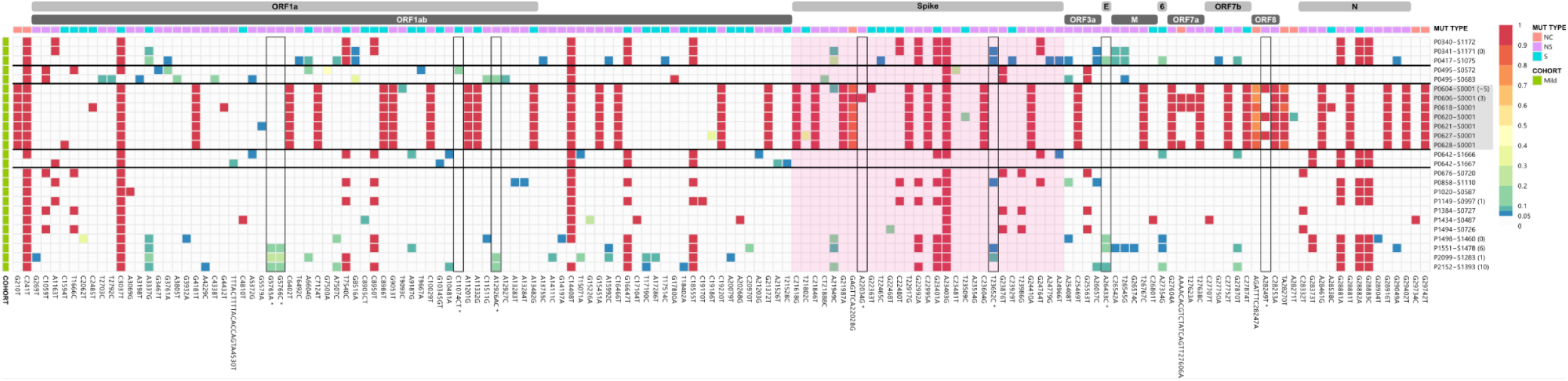
iSNV and SNP frequency changes and the corresponding synonymous and non-synonymous mutations across the SAR-CoV-2 genomes for the mild clinical cohort. A frequency of ≥ 0.9 was considered a SNP (red), frequencies below 0.05 were not included in the analysis. Identified problematic sites are outlined in black and denoted with an * on the x axis. The spike gene is highlighted in pink.

**Supplementary Figure S3:**
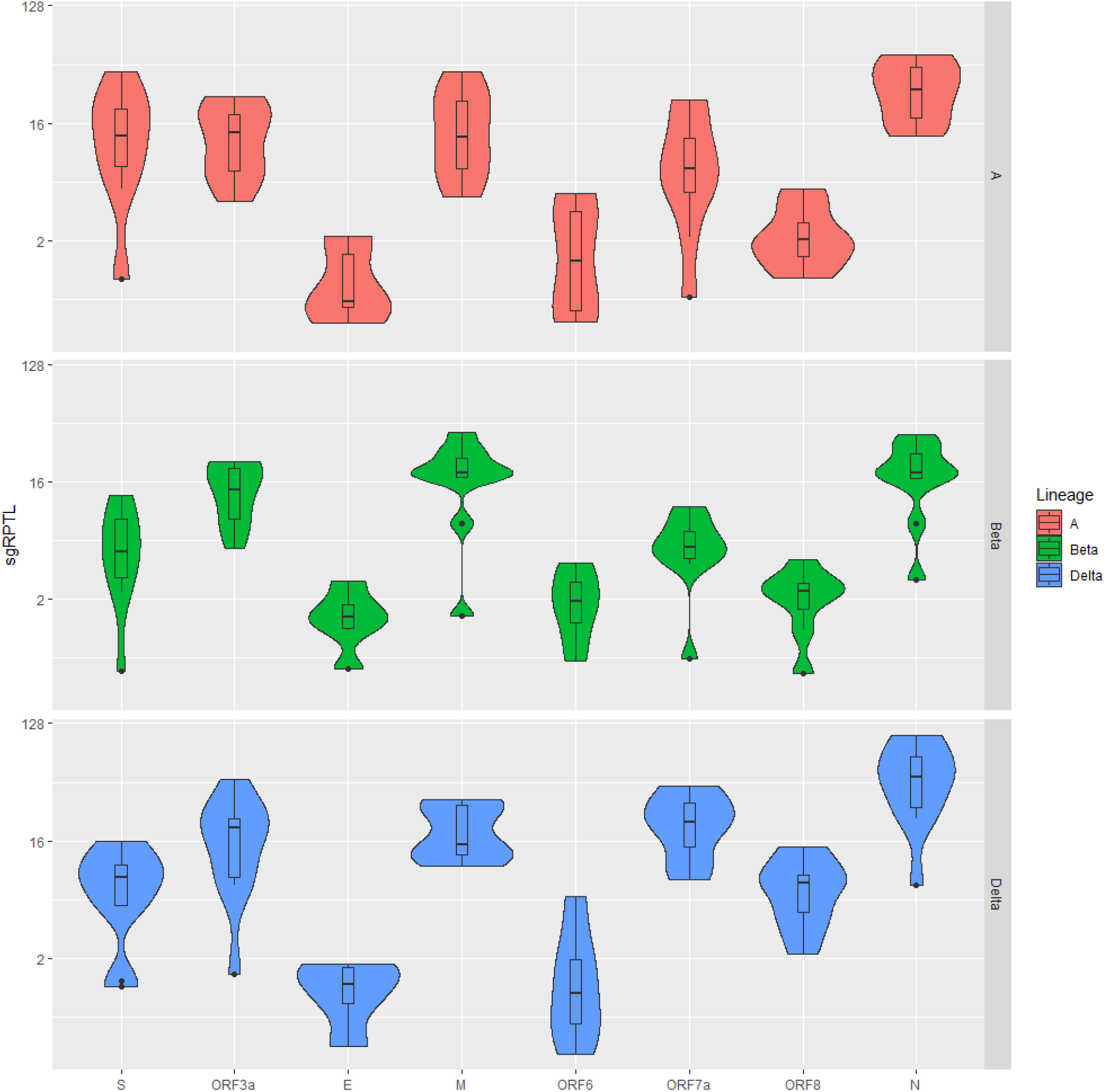
Violin plots depicting the sgRPTL normalized counts for sgRNA abundance at each gene from top to bottom of Culture A, Culture Beta, and Culture Delta. There was significantly higher sgRNA across all genes in Delta compared to A and Beta. A and Beta were not significantly different. Individually, N was significantly higher in Delta compared to Beta and ORF 7a and 8 were significantly high than both A and Beta. The boxplots within the violins indicate the median and the interquartile ranges (25th to 75th percentile).

